# Race-adjusted Lung Function Increases Inequities in Diagnosis and Prognosis and Should Be Abandoned

**DOI:** 10.1101/2022.01.18.22269455

**Authors:** Magnus Ekström, David Mannino

## Abstract

**Background:** Lung function assessment is essential for respiratory medicine and health. Recommended international reference values differ by race, which is controversial. We evaluated the effect of adjusting lung function for race on prevalence of lung function impairment, breathlessness and mortality in the US population.

**Methods:** Population-based analysis of the National Health and Nutrition Examination Survey (NHANES) 2007–2012. Race was analyzed as black, white, or other. Lung function was assessed as forced expired volume in one second (FEV_1_) and forced vital capacity (FVC). Predicted normal values were calculated for each person using the Global Lung Initiative (GLI)-2012 equations for 1) white; 2) black; and 3) other/mixed populations. Outcomes were compared for the different reference values in relation to: prevalence of lung function impairment (<lower limit of normal [LLN]), moderate/severe impairment (<50%pred); self-reported exertional breathlessness; and mortality up to 31 December, 2015.

**Findings:** We studied 14,123 people (50% female); white (n=5,928), black (n=3,130), and other (n=5,065). Compared to those for white, black reference values identified markedly fewer cases of lung function impairment (FEV_1_) both in black people (9.3% vs. 36.9%) and other non-white races (1.5% vs. 9.5%); and prevalence of moderate/severe impairment was approximately halved. Outcomes among those impaired differed by reference value used: white (best outcomes), other/mixed (intermediate), and black (worst outcomes). Black people with FEV_1_ ≥LLN_black_ but <LLN_white_ had 48% increased rate of breathlessness and almost doubled mortality, compared to blacks ≥LLN_white_. Lung function ≥LLN_white_ identified people with good outcomes, similarly in black and white people. Findings were similar when analyzing FEV_1_ or FVC.

**Interpretation:** Race adjustment of lung function should be abandoned. White reference values are most sensitive and specific to identify impairment, and could be applied across the population for improved assessment and health equity.

**Funding:** Swedish Research Council (Dnr: 2019-02081).

**Research in context:** *Evidence before this study:* We searched MEDLINE and Embase using search terms including “race”, “ethnicity”, “pulmonary function”, “spirometry”, and “prediction equations” from database inception and January 10, 2022, for papers published in English. A total 33 papers related to lung function and race were identified. Race-adjusted lung function reference values were recommended by major guidelines for use internationally. Race-specific references assume a 10-15% lower lung function, such as the forced expired volume in one second (FEV_1_) and forced vital capacity (FVC), in black people and 4-6% lower in Asian people compared with in whites. Compared to not adjusting for race, race-adjusted lung function values have recently been questioned as they have been found to not improve prediction of outcomes in population-based studies or in people at risk of obstructive pulmonary disease. Concerns have been raised that, contrary to the intent, race-adjusted reference values may contribute to under diagnosis of disease in disadvantage minorities, with the largest differences reported in black (Afro-American) people, and may worsen race-related health inequalities. Data on the impact of race-adjusted lung function values across the ethnically diverse population are limited and data on how to decrease racial bias in lung function assessment are needed.

*Added value of this study:* We analyzed the impact of using different race-specific (GLI-2012) reference equations for FEV_1_ and FVC across the US population in the National Health and Nutrition Examination Survey (NHANES) 2007-2012. Outcomes were prevalence of lung function impairment (value < lower limit of normal), breathlessness on exertion, and mortality up to December 31, 2015. Compared to using references for whites, black reference values were less likely to identify lung function impairment across all races but especially in blacks (9.3% vs. 36.9%); and those identified had lower lung function, more breathlessness, and worse prognosis. Black people with lung function normal by black standards but impaired by white standards had increased prevalence of breathlessness and mortality, compared to those normal also by white standards. Thus, race-adjusted reference values labeled black people as normal despite worse outcomes. White normal values identified people with similarly good lung function, and low rates of breathlessness and mortality across races groups.

*Implications of all the available evidence:* The findings from this study support that race-adjusted reference values markedly under diagnose lung function impairment, and related breathlessness, and mortality in underprivileged groups across the US population. Normal values for whites were most sensitive to identify lung function impairment related to worsening outcomes and people classified as having normal lung function with similar good outcomes irrespective of race group. These findings suggest that lung function should not be adjusted for race. When applied across the population, white reference values were most sensitive to identify smaller or earlier impairment and most specific to identify people with normal lung function with similarly good outcomes across race groups. Given the large impact shown, abandoning the use of race-adjusted lung function values is likely to contribute to improved health equity.

## Introduction

Chronic respiratory disease is the third leading cause of death worldwide.^1^ Pulmonary function testing using spirometry is key to the diagnosis of chronic respiratory disease, evaluation of breathlessness, whether people qualify for interventions such as lung transplant, or can be considered to be disabled.^2^ Standards exist for both the performance ^3^ and interpretation ^4^ of spirometry. The American Thoracic Society (ATS) recommends that ‘laboratories must select appropriate reference values for the patients being tested’ ^4^ and goes on to recommend use of the Global Lung Initiative (GLI)-2012 prediction equations,^5^ which establishes race–specific reference values for whites, African Americans, North East Asians, and South East Asians. Currently, race–specific reference values for lung function are the recommended standard for use internationally.^6-8^

How do race–specific reference values for lung function work? The GLI-2012 prediction equations for normal lung function adjust for age, height, sex, and race.^3^ While historic prediction equations would apply an ‘adjustment factor’ of 0.88 (12% less) for black populations and 0.94 (6% less) for Asian populations,^8^ the GLI-2012 equations were developed without a fixed adjustment factor but rather using race-specific populations. However, even in the GLI-2012 equations, predicted lung function levels are 10-15% lower in African Americans and South East Asians relative to whites and North East Asians.^2^

Race adjustment is controversial. On one side of the argument is the thought that race is a surrogate measure that captures a number of factors predictive of poor health status and outcomes that are not really specific to a person’s racial make-up.^9^ The other side of the argument is that there are physiologic traits between populations that are based in genetics and captured, to some extent, by self-reported race.^10^ This is analogous to the differences between men and women in predicted lung function, even with identical heights.^11^ In other area of medicine, race-specific normal values have recently been shown to discriminate and contribute to under-diagnosing and under-treatment in socioeconomically more vulnerable groups such as Afro-Americans and are currently revised not to adjust for race, such as normal values for renal function.^12^

We aimed to determine the effect of adjusting lung function for race on prevalence of lung function impairment, breathlessness and mortality in the US population.

## Methods

### Design and population

This was a population-based analysis of the National Health and Nutrition Examination Survey (NHANES) in the US from 2007 to 2012.^13-15^ We included all people aged ≥ 18 years with data on demographics and spirometry. Data were obtained on breathlessness on exertion (available for people aged ≥ 40 years), and mortality up to 31 December 2015. Participants provided written consent to participate in NHANES using a protocol approved by the National Center for Health Statistics Research Ethics Review Board.^13-15^ The present analysis used only de-identified NHANES data which are publicly available. The study is reported in accordance with STrengthening the Reporting of OBservational studies in Epidemiology (STROBE) guidelines.^16^

### Assessments

Data on age, sex, and race were from personal interviews. Race was categorized as Non-Hispanic whites (whites), Non-Hispanic black (blacks), and others (Hispanic, Asian, mixed race, etc.). Analysis focused on comparing black *vs*. white as these reference values differ the most.^5^ The category other was included to reflect the entire NHANES population.

Measured weight (kg), height (cm), and spirometry were obtained using mobile examination centres. Dynamic spirometry was performed in accordance with guidelines from the ATS and European Respiratory Society (ERS).^17^ Values were recorded as the highest obtained value (pre- or post-bronchodilator). For each participant, reference values for the forced expired volume in one second (FEV_1_) and forced vital capacity (FVC) were calculated using the GLI-2012 equations for 1) white, 2) black, and 3) other/mixed populations.^5^ Thus, for each individual we calculated three predicted reference values for FEV_1_ and FVC, respectively, for comparison.

Breathlessness on exertion was assessed using the question: ‘Have you had shortness of breath either when hurrying on the level or walking up a slight hill?’ (yes/no), corresponding to a breathlessness level of ≥ 1 point on the modified Medical Research Council (mMRC) scale.^18^ Breathlessness data were available in NHANES for people 40 years or older. Mortality was assessed using standardized NHANES procedures up to 31 December 2015.

### Statistical analyses

The study population was weighted (using published NHANES weights for people undergoing examinations including spirometry), to represent the non-institutionalized US population during the six year period. For all analyses, variance estimates were produced using Taylor Series Linearization methods,^19^ as recommended for NHANES.

Data were tabulated and compared between race groups using means (standard deviation [SD]) for normally distributed continuous variables, and frequency (percentage) for categorical variables. Lung function was evaluated as FEV_1_ in the main analyses. Similar analyses of FVC are reported in the supplements.

Outcomes were compared between race groups (white, black, and other) in terms of: 1) predicted normal values using each race-specific prediction equation (white, black, or other/mixed); 2) prevalence of impaired lung function, defined as value < the lower limit of normal (LLN) using each race-specific prediction equation; and the prevalence of moderate to severe impairment, defined as < 50% of the predicted normal in accordance with GOLD (Global Initiative for Obstructive Lung Disease);^6^ 3) prevalence of breathlessness; and 4) mortality. As each prediction equation was applied to the same people, the effects of applying different race– specific reference values were independent of (adjusted for) participant characteristics by design.

Breathlessness and mortality were compared by race and lung function impairment (defined using different race-specific prediction equations) using five mutually exclusive categories: ‘White Normal’ (white race, value ≥ LLN_white_); ‘White Abnormal’ (white race, value < LLN_white_); ‘Black Normal’ (white race, value ≥ LLN_white_); ‘Black Abnormal (White Standard)’ (black race, value <LLN_white_ but ≥ LLN_black_); or ‘Black Abnormal (Black Standard)’ (black race and value < LLN_white_ and <LLN_black_). As normal values were higher for all persons using white than black prediction equations, all values < LLN_black_ were also <LLN_white_. Probability of breathlessness was analyzed using logistic regression and was expressed as relative rate ratios (RRR). Mortality was analyzed using Cox proportional-hazards regression and expressed as hazard ratios (HR). Associations with breathlessness and mortality were also analyzed for lung function impairment using each race –specific prediction equation in the whole population, adjusting for age, sex and body mass index (BMI). All estimates were reported with 95% confidence intervals (CIs). Statistical analyses were performed with Stata version 16.0 (StataCorp LP; College Station, TX).

## Results

A total 14,123 people (50% female) were studied with race self-reported as white (n=5,928), black (n=3,130), or other (n=5,065). Compared with the other groups, white people were slightly older and had somewhat higher absolute FEV_1_ and FVC values, whereas the sex distribution and BMI was similar between race groups (Table 1). Breathlessness was more prevalent among black people, as compared with white and other (Table 1; p < 0.001 for both comparisons).

**Table 1.** Characteristics and lung function by race

| Factor | White people | Black people | Other |
| --- | --- | --- | --- |
| N | 5,928 | 3,130 | 5,065 |
| Age, mean (SD) | 45.7 (16.0) | 42.1 (15.8) | 39.5 (14.5) |
| Female, % | 50.3% | 53.7% | 48.3% |
| Weight (kg), mean (SD) | 83.0 (20.7) | 87.9 (24.0) | 76.7 (19.2) |
| Height (cm), mean (SD) | 170.7 (9.8) | 169.6 (9.5) | 165.1 (9.6) |
| Body mass index (kg/m<sup>2</sup>), mean (SD) | 28.4 (6.5) | 30.6 (8.1) | 28.1 (6.2) |
| FEV<sub>1</sub> (L), mean (SD) | 3.3 (0.9) | 2.9 (0.8) | 3.2 (0.8) |
| FVC (L), mean (SD) | 4.3 (1.1) | 3.6 (1.0) | 4.0 (1.0) |
| FEV<sub>1</sub>/FVC, mean (SD) | 0.77 (0.08) | 0.80 (0.08) | 0.81 (0.07) |
| Breathlessness prevalence*, % | 29.3% | 33.7% | 24.4% |
| Deaths by December 31, 2015, % | 3.0% | 3.5% | 2.2% |

Characteristics are weighted to represent the mean US population in National Health and Nutrition Examination Survey (NHANES) 2007-2012. \* Breathlessness prevalence is in people aged 40 years or older. Abbreviations: FEV<sub>1</sub> = forced expired volume in one second. FVC = forced vital capacity.

### Prevalence of lung function impairment

The predicted normal FEV_1_ was highest using the equation for whites, intermediate using that for other/mixed, and lowest when using the equation for black people (Table 2). This pattern was similar in all race groups. In black people the predicted normal FEV_1_ dropped from 3.5 liter using the white equation to 3.0 liter using the black equation, a decrease by 14%.

**Table 2.**
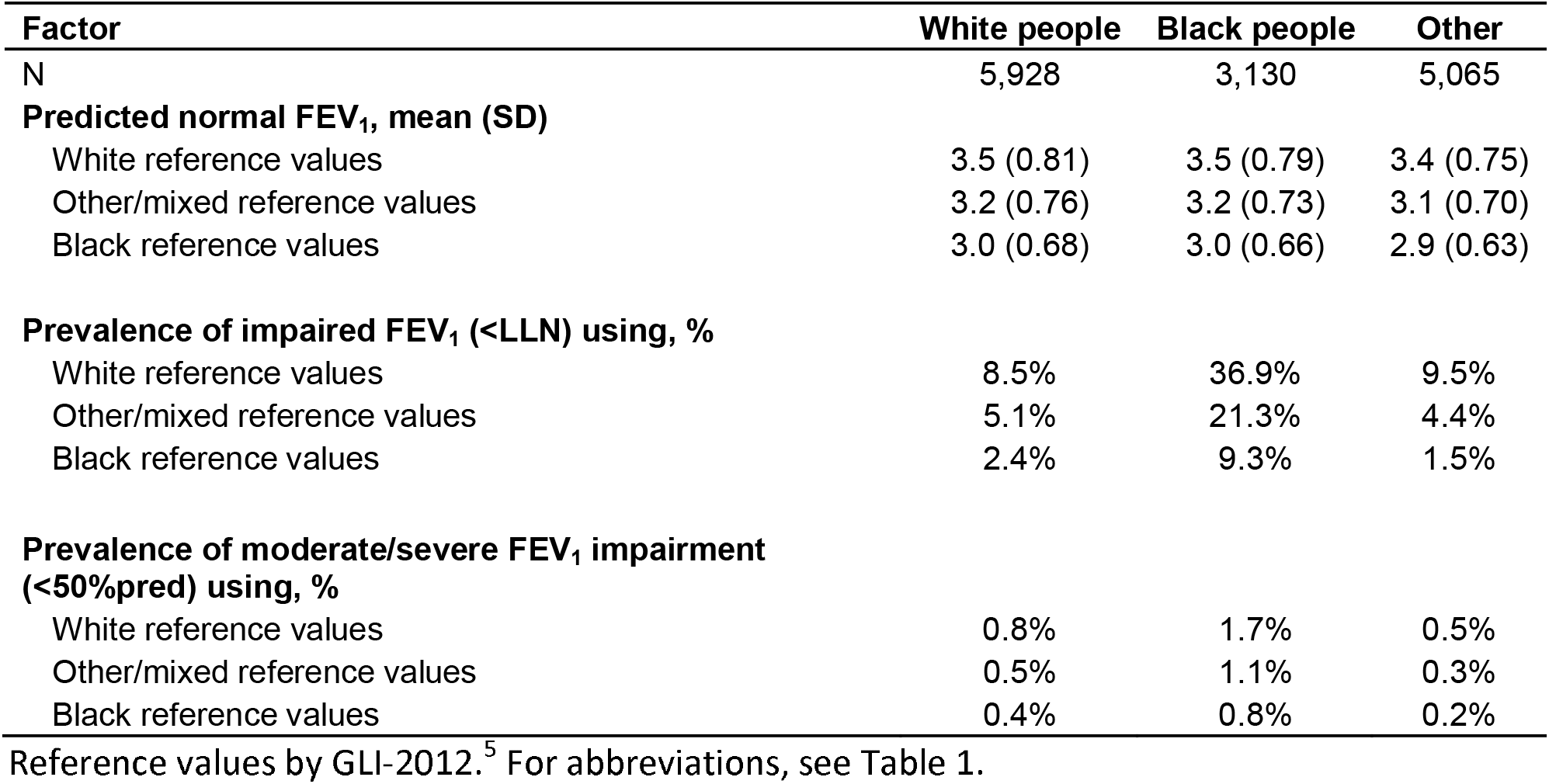
Prevalence of impaired FEV_1_ by race and race-specific reference value used

| **Factor** | **White people** | **Black people** | **Other** |
| --- | --- | --- | --- |
| N | 5,928 | 3,130 | 5,065 |
| **Predicted normal FEV<sub>1</sub>, mean (SD)** |  |  |  |
| White reference values | 3.5 (0.81) | 3.5 (0.79) | 3.4 (0.75) |
| Other/mixed reference values | 3.2 (0.76) | 3.2 (0.73) | 3.1 (0.70) |
| Black reference values | 3.0 (0.68) | 3.0 (0.66) | 2.9 (0.63) |
| **Prevalence of impaired FEV<sub>1</sub> (<LLN) using, %** |  |  |  |
| White reference values | 8.5% | 36.9% | 9.5% |
| Other/mixed reference values | 5.1% | 21.3% | 4.4% |
| Black reference values | 2.4% | 9.3% | 1.5% |
| **Prevalence of moderate/severe FEV<sub>1</sub> impairment (<50%pred) using, %** |  |  |  |
| White reference values | 0.8% | 1.7% | 0.5% |
| Other/mixed reference values | 0.5% | 1.1% | 0.3% |
| Black reference values | 0.4% | 0.8% | 0.2% |

Reference values by GLI-2012.<sup>5</sup> For abbreviations, see Table 1.

The choice of race adjustment strongly influenced the prevalence of lung function impairment (Table 2). Compared with the equation for white people, black reference values identified markedly fewer cases of impaired lung function, both among whites (2.4% vs. 8.5%), in black people who had the largest absolute decrease (9.3% vs. 36.9%), as well as in other races (1.5% vs. 9.5%). Overall, compared with the reference values for blacks, white reference values identified about four times as many people as having impaired lung function, and identification of moderate to severe impairment (< 50% predicted) was approximately doubled (Table 2). All findings were similar when analyzing FVC instead of FEV_1_ (supplemental Table S1).

### Lung function and outcomes

People with impaired lung function (FEV_1_<LLN) had increased rates of breathlessness and mortality across the whole population, but outcomes differed by the race-specific reference value used: black (worst outcomes), other/mixed (intermediate), and white (best outcomes). Breathlessness associations using each race–specific reference value were: for black (RRR 4.6; 95% CI, 3.2 – 6.6), other/mixed (RRR 3.4; 95% CI, 2.7 – 4.4), and white (RRR 2.8; 95% CI, 2.4 – 3.3). Mortality estimates were: for black (HR 3.5; 95% CI, 2.4 – 5.2), other/mixed (HR 2.8; 95% CI, 2.1 – 3.6), and white (HR 2.6; 95% CI, 2.1 – 3.4).

Outcomes by race (black or white) and lung function impairment defined using the different race–specific reference values are shown in Figure 1. People with normal lung function according to white standards (≥ LLN_white_) had low rates or breathlessness and mortality which were similar in both white and black people. In contrast, black reference values introduced a race–related bias; black people with lung function normal by the black standard but impaired by the standard for whites (FEV_1_ ≥ LLN_black_ but < LLN_white_) had significantly increased rates of breathlessness (RRR 1.48; 95% CI, 1.13–1.94) and mortality (HR 1.87; 95% CI, 1.42–2.46), compared to black people who were also normal by white standards (FEV_1_ ≥ LLN_white_). Associations from Cox regression are shown in supplemental Table S2.

**Figure 1.**
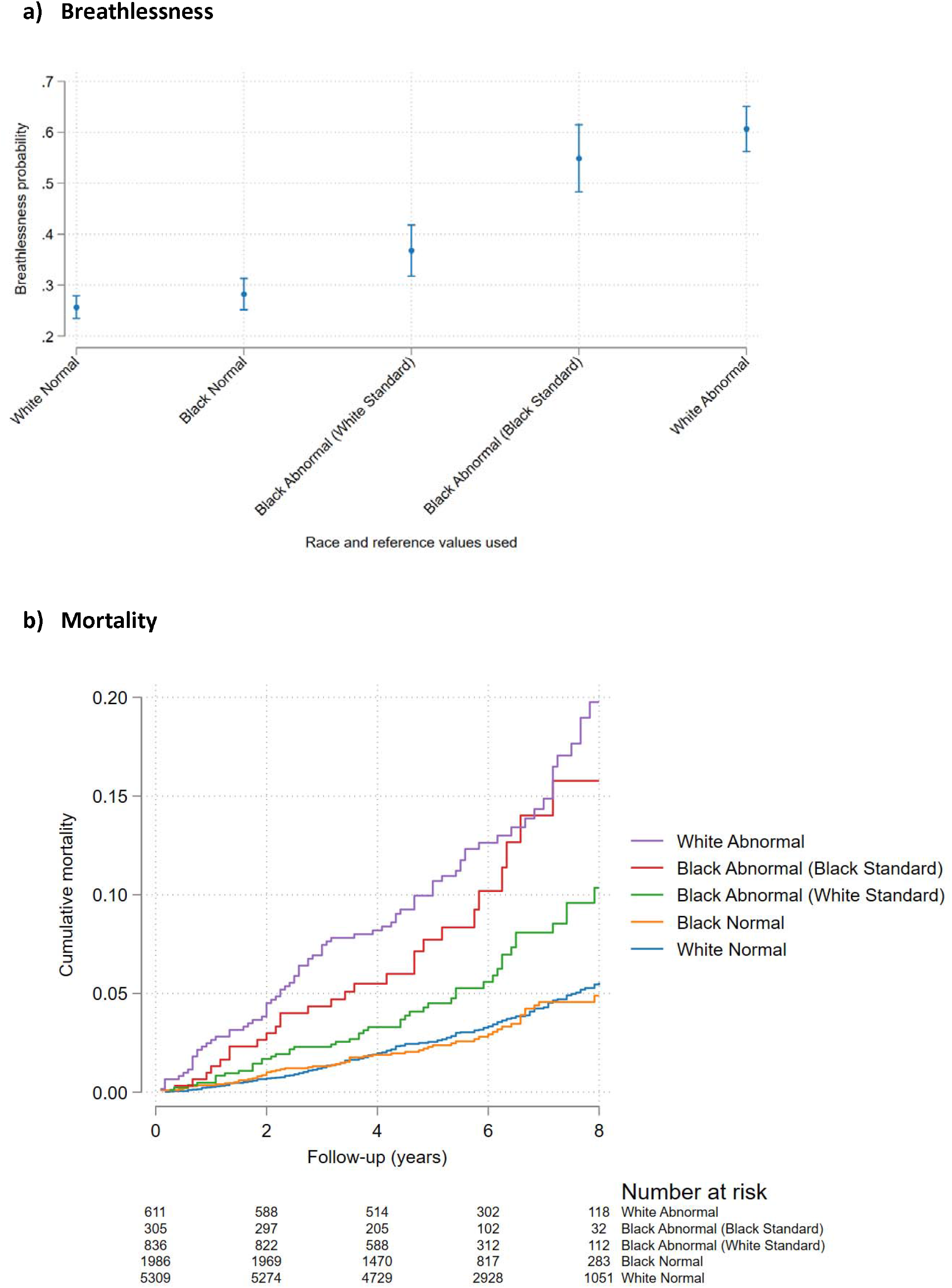
Outcomes by race and FEV_1_ impairment defined using white and/or black normal values, in terms of a) breathlessness, and b) mortality. Breathlessness probability was analyzed using logistic regression, and mortality using Cox proportional hazards regression. Impaired lung function was defined as a forced expired volume in one second (FEV_1_) < lower limit of normal (LLN) using GLI-2012 predicted normal values for white and black people, respectively.^5^ Groups were categorized by race and FEV_1_ impairment according to different race-specific prediction equations as: ‘White Normal’ (white race with FEV_1_ ≥ LLN_white_); ‘Black Normal’ (black race with FEV_1_ ≥ LLN_white_); ‘Black Abnormal (White Reference)’ (black race with FEV_1_<LLN_white_ but ≥ LLN_black_); ‘Black Abnormal (Black Reference)’ (black race and FEV_1_ < LLN_white_ and <LLN_black_); and ‘White Abnormal’ (white race and FEV_1_<LLN_white_). The main finding is that black people who were categorized as having a normal FEV_1_ using LLN_black_ but not using LLN_white_ had increased breathlessness prevalence and mortality compared with people categorized as normal using reference values for white. Thus, black reference values misclassify black people as having normal lung function despite having worse outcomes. When defining normality using LLN_white_ for all, people with normal FEV_1_ had similar breathlessness and mortality in both white and black people.

Findings were similar when analyzing FVC (Table S3 and Figure S1 in the supplement).

## Discussion

The main findings are that, compared with using reference values (GLI-2012)^5^ for white people, race-specific equations for black or mixed/other populations markedly under diagnose lung function impairment, including moderate to severe disease, and misclassify black people as having normal lung function despite having substantially increased rates of breathlessness and mortality. In black people, race adjusted (black) reference values identified only ¼ of cases (9.3% *vs*. 36.9%) of impaired FEV_1_ as compared with white reference values – and those identified by the black references had significantly worse lung function, more breathlessness and increased mortality. Black people with normal lung function according to black reference values but impaired according to those for whites had 48% increased rate of breathlessness and almost doubled mortality, as compared with black people with normal FEV_1_ by white standards. This racial bias was largely avoided by applying the white reference values across the whole population; besides identifying the higher prevalence of lung function impairment with worse outcomes in black, people with normal lung function according to white reference values had similar good prognosis across all the race groups.

These findings have important implications for assessment of lung function and respiratory disease. Firstly, use of race-adjusted reference values, which are currently endorsed by major international guidelines,^6, 7^ should be abandoned. As we show, race-adjusted references (compared to white reference values) misclassify lung function as normal despite worse outcomes in as many as 28% of black people, corresponding to as many as 13.1 million people in the US alone.^20^ A particularly alarming finding was that half of cases of moderate to severe lung function impairment were missed – which could lead to insufficient treatment or delayed interventions such as lung transplantation evaluation. Adjusting lung function values for race thus contribute to under diagnosis of disease and disability, failure to identify impaired lung function as contributing to breathlessness, potentially leading to insufficient or delayed treatment and compounded race-related health inequities. Second, to avoid this race-related bias, the present findings support that lung function should be assessed using a common prediction equation across the population. The choice of prediction equation may vary depending on aim of the assessment. In research and clinical medicine, the aims of spirometry are mainly to evaluate whether lung function is normal or impaired, whether breathlessness is related to impaired lung function, evaluate the severity of respiratory disease needing treatment or further evaluation, and to predict prognosis. With emphasis on sensitivity to detect and treat lung function impairment earlier, the present findings support the use of the GLI-2012 prediction equations for white people across the population. We found white references to be more sensitive than the GLI-2012 mixed/other reference values, which were previously proposed for use across mixed populations.^5^ Importantly, while being more sensitive, white reference predictions were still specific to identify impairment associated with substantial morbidity in terms of increased breathlessness and mortality rates, which was seen across all the race groups. White reference values identified smaller or earlier lung function impairment, which should be appropriately evaluated, and may be more amenable to treatment.

The present findings extend previous reports that race-related differences in mortality were attenuated by applying using the same prediction equation (reference values for whites) across the population.^21^ The findings are in line with those that using race-adjusted reference values (compared to not adjusting for race) did not improve prediction of respiratory morbidity or mortality in a large cohort study,^22^ and predicted clinically important outcomes worse in black and white people at high risk of COPD (n=2,652).^23^ Taken together, these previous data support our findings that using race-specific reference values, rather than counteract, in fact widen race-related inequities.

Lower lung function in black people is consistent with data that airflow obstruction and reduced lung function strongly associates with poverty at individual and community levels across multiple countries, independent of factors such as age, sex, and smoking and tuberculosis.^24^ In the recent study by Baugh *et al*., controlling for comorbid disease and measures of adversity weakened the association between race and FEV_1_, suggesting that racial differences in lung function are at least partly reflect differential exposures.^23^ It is increasingly acknowledged that race is, to a large part, a social construct.^2^ Genetic and environmental factors inseparably interact in multiple and complex ways to influence all aspects of life, through prenatal and early life factors, circumstances throughout life, over the generations.^2^ In the Eight America’s project, Murray *et al*. described large disparities in mortality across race-county groupings and concluded that these differences could not be explained by race, income, or basic health-care access and utilization alone.^25^ As pointed out,^2^ the lower lung function in disadvantaged groups including Afro-Americans might, to an extent, reflect a higher accumulated exposure to adverse exposures and not disease. But as we show, race-adjusted lung function values may obscure the higher prevalence of impairment in these populations, misclassify people as healthy despite having worse outcomes, and contribute to under diagnosis of disease or presence of modifiable health exposures that could, when appreciated, be modified.^2^ An example of the influence of environmental factors from another field is the generational change in health outcomes when comparing the population of southern Europe to northern Europe.^26^ Even though these populations had similar race distributions, large differences in both adult height and childhood mortality seen in 1950 had largely disappeared by 1980.^26^

The suggestion that race-adjustment should be abandoned in lung function assessment is consistent with similar developments to counteract racial bias in other medical areas, including tests in haematology, and references for kidney function.^12, 27, 28^

Strengths of the present study include the use of a well characterized, large database representative for the racially diverse non-institutionalized US population. Race-specific prediction equations for normal lung function (FEV_1_ and FVC) were evaluated using the international GLI-2012 reference values developed to be applicable globally, in accordance with guidelines.^5-7^ By comparing the predictions in the same population, the analyses were independent of differences in participant characteristics. Reference values were evaluated against clinically important outcomes in terms of prevalence of impairment, breathlessness and mortality.

A limitation of the present study is that data pertain to the US population, and studies in other settings are needed. However, we show how currently recommended race-specific prediction equations for lung function perform, introduce race-related bias, and now this bias may be attenuated or overcome by applying a similar reference across a large diverse population. The findings are likely to be valid and relevant for avoiding race–related bias in lung function assessment in many settings around the world.

In conclusion, reference values for lung function should not be adjusted for race, as it leads to substantial under diagnosis of impairment and misclassification of health status and outcomes in under privileged groups. Lung function should be evaluated using a common prediction equation across races, where reference values for whites are most sensitive to detect smaller and earlier impairment, which could promote improved management and health equity.

## Data Availability

All data used in the analyses are publicly available at: https://www.cdc.gov/nchs/nhanes/index.htm

## Contributors statement

Both authors conceptualized the study design; ME performed the analyses, and wrote the first draft. Both authors verified the data and analyses, and were involved in the interpretation, revision and decision to submit the final manuscript.

## Data sharing statement

All data used are anonymized and publicly available, and scripts (for Stata) of all analyses will be made available from the authors upon reasonable request.

## Supplementary material

**Table S1.**
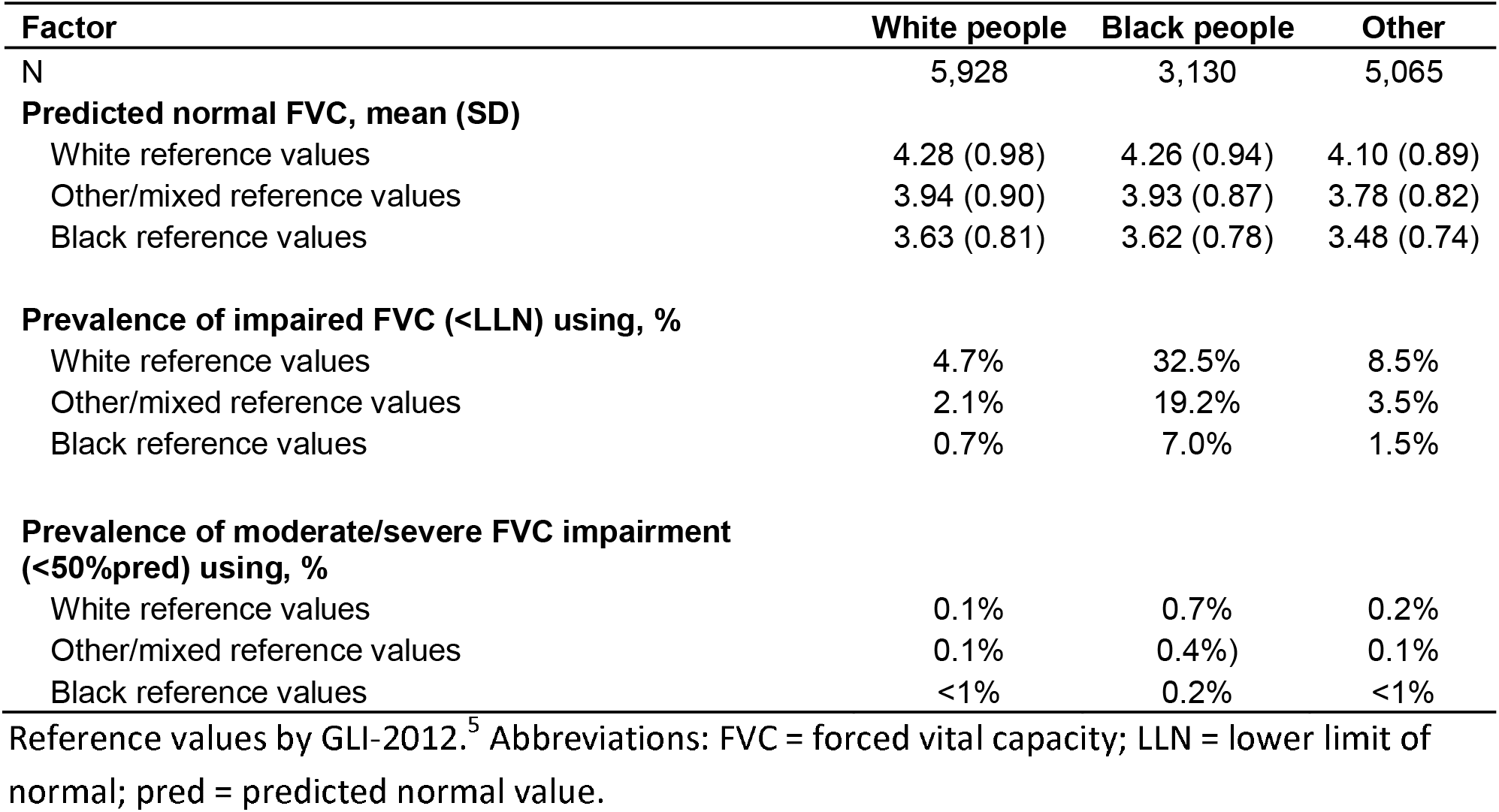
Prevalence of impaired FVC by race and race-specific reference values used

**Table S2.**
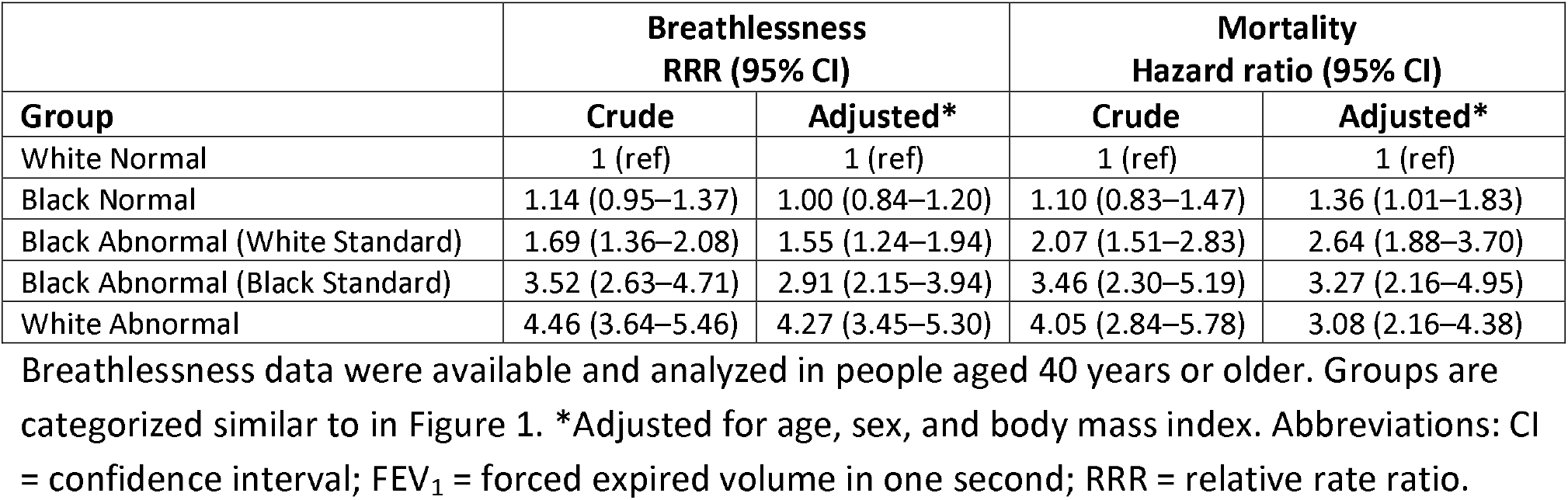
Associations with breathlessness and mortality by race and FEV_1_ impairment

**Table S3.**
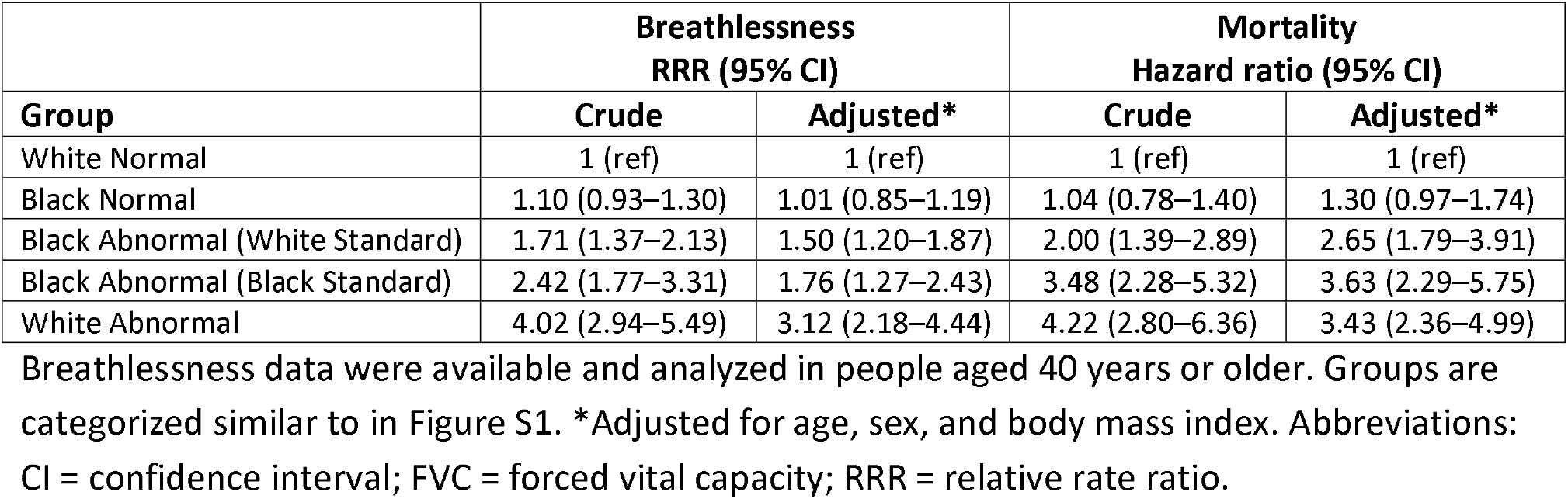
Associations with breathlessness and mortality by race and FVC impairment

**Figure S1.**
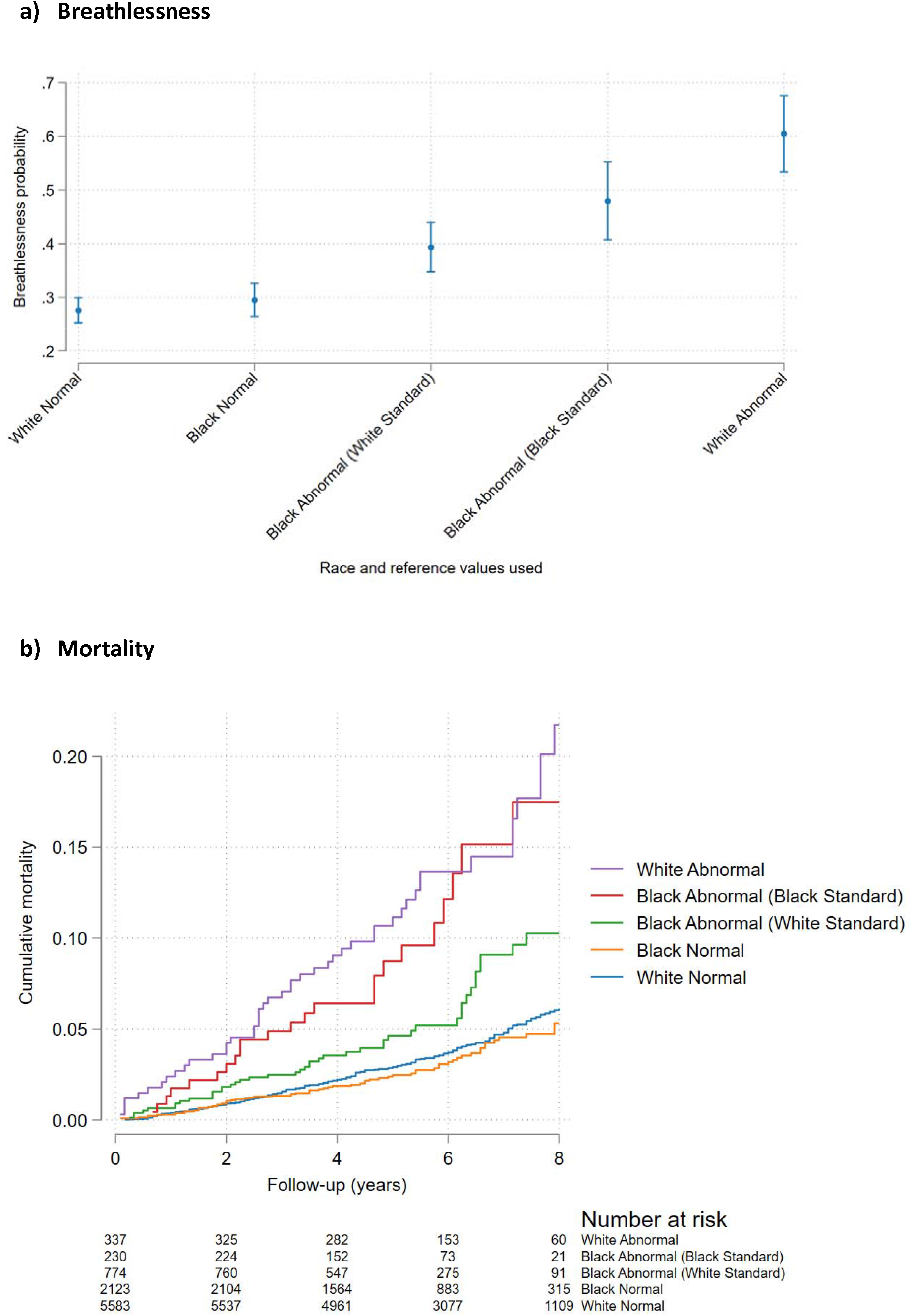
Outcomes by race and FVC impairment defined using reference values for white and/or black people, in terms of a) breathlessness, and b) mortality. Breathlessness probability was analyzed using logistic regression, and mortality using Cox proportional hazards regression. Impaired lung function was defined as a forced vital capacity (FVC) < lower limit of normal (LLN) using GLI-2012 predicted normal values for white and black people, respectively.^5^ Groups were categorized by race and FVC impairment according to different race-specific prediction equations as: ‘White Normal’ (white race with FVC ≥ LLN_white_); ‘Black Normal’ (black race with FVC ≥ LLN_white_); ‘Black Abnormal (White Reference)’ (black race with FVC < LLN_white_ but ≥ LLN_black_); ‘Black Abnormal (Black Reference)’ (black race and FVC < LLN_white_ and <LLN_black_); and ‘White Abnormal’ (white race and FVC < LLN_white_). The main finding is that black people who were categorized as having a normal FVC using LLN_black_ but not using LLN_white_ had increased breathlessness prevalence and mortality compared with people categorized as normal using reference values for white. Thus, black reference values misclassify black people as having normal lung function despite having worse outcomes. When defining normality using LLN_white_ for all, people with normal FVC had similar breathlessness and mortality in both white and black people.

