## Supplemental material for "Race-adjusted Lung Function Increases Inequities in Diagnosis and Prognosis and Should Be Abandoned"

**Table S1.** Prevalence of impaired FVC by race and race-specific reference values used

| **Factor** | **White people** | **Black people** | **Other** |
| --- | --- | --- | --- |
| N | 5,928 | 3,130 | 5,065 |
| **Predicted normal FVC, mean (SD)** |  |  |  |
| White reference values | 4.28 (0.98) | 4.26 (0.94) | 4.10 (0.89) |
| Other/mixed reference values | 3.94 (0.90) | 3.93 (0.87) | 3.78 (0.82) |
| Black reference values | 3.63 (0.81) | 3.62 (0.78) | 3.48 (0.74) |
| **Prevalence of impaired FVC (<LLN) using, %** |  |  |  |
| White reference values | 4.7% | 32.5% | 8.5% |
| Other/mixed reference values | 2.1% | 19.2% | 3.5% |
| Black reference values | 0.7% | 7.0% | 1.5% |
| **Prevalence of moderate/severe FVC impairment (<50%pred) using, %** |  |  |  |
| White reference values | 0.1% | 0.7% | 0.2% |
| Other/mixed reference values | 0.1% | 0.4%) | 0.1% |
| Black reference values | <1% | 0.2% | <1% |

Reference values by GLI-2012.^5^ Abbreviations: FVC = forced vital capacity; LLN = lower limit of normal; pred = predicted normal value.

**Table S2.** Associations with breathlessness and mortality by race and FEV_1_ impairment

|  | **Breathlessness**  **RRR (95% CI)** | | **Mortality**  **Hazard ratio (95% CI)** | |
| --- | --- | --- | --- | --- |
| **Group** | **Crude** | **Adjusted*** | **Crude** | **Adjusted*** |
| White Normal | 1 (ref) | 1 (ref) | 1 (ref) | 1 (ref) |
| Black Normal | 1.14 (0.95–1.37) | 1.00 (0.84–1.20) | 1.10 (0.83–1.47) | 1.36 (1.01–1.83) |
| Black Abnormal (White Standard) | 1.69 (1.36–2.08) | 1.55 (1.24–1.94) | 2.07 (1.51–2.83) | 2.64 (1.88–3.70) |
| Black Abnormal (Black Standard) | 3.52 (2.63–4.71) | 2.91 (2.15–3.94) | 3.46 (2.30–5.19) | 3.27 (2.16–4.95) |
| White Abnormal | 4.46 (3.64–5.46) | 4.27 (3.45–5.30) | 4.05 (2.84–5.78) | 3.08 (2.16–4.38) |

Breathlessness data were available and analyzed in people aged 40 years or older. Groups are categorized similar to in Figure 1. *Adjusted for age, sex, and body mass index. Abbreviations: CI = confidence interval; FEV_1_ = forced expired volume in one second; RRR = relative rate ratio.

**Table S3.** Associations with breathlessness and mortality by race and FVC impairment

|  | **Breathlessness**  **RRR (95% CI)** | | **Mortality**  **Hazard ratio (95% CI)** | |
| --- | --- | --- | --- | --- |
| **Group** | **Crude** | **Adjusted*** | **Crude** | **Adjusted*** |
| White Normal | 1 (ref) | 1 (ref) | 1 (ref) | 1 (ref) |
| Black Normal | 1.10 (0.93–1.30) | 1.01 (0.85–1.19) | 1.04 (0.78–1.40) | 1.30 (0.97–1.74) |
| Black Abnormal (White Standard) | 1.71 (1.37–2.13) | 1.50 (1.20–1.87) | 2.00 (1.39–2.89) | 2.65 (1.79–3.91) |
| Black Abnormal (Black Standard) | 2.42 (1.77–3.31) | 1.76 (1.27–2.43) | 3.48 (2.28–5.32) | 3.63 (2.29–5.75) |
| White Abnormal | 4.02 (2.94–5.49) | 3.12 (2.18–4.44) | 4.22 (2.80–6.36) | 3.43 (2.36–4.99) |

Breathlessness data were available and analyzed in people aged 40 years or older. Groups are categorized similar to in Figure S1. *Adjusted for age, sex, and body mass index. Abbreviations: CI = confidence interval; FVC = forced vital capacity; RRR = relative rate ratio.

**Figure S1.**  **Outcomes by race and FVC impairment defined using reference values for white and/or black people, in terms of a) breathlessness, and b) mortality.** Breathlessness probability was analyzed using logistic regression, and mortality using Cox proportional hazards regression. Impaired lung function was defined as a forced vital capacity (FVC) < lower limit of normal (LLN) using GLI-2012 predicted normal values for white and black people, respectively.^5^ Groups were categorized by race and FVC impairment according to different race-specific prediction equations as: ‘White Normal’ (white race with FVC ≥ LLN_white_); ‘Black Normal’ (black race with FVC ≥ LLN_white_); ‘Black Abnormal (White Reference)’ (black race with FVC < LLN_white_ but ≥ LLN_black_); ‘Black Abnormal (Black Reference)’ (black race and FVC < LLN_white_ and <LLN_black_); and ‘White Abnormal’ (white race and FVC < LLN_white_). The main finding is that black people who were categorized as having a normal FVC using LLN_black_ but not using LLN_white_ had increased breathlessness prevalence and mortality compared with people categorized as normal using reference values for white. Thus, black reference values misclassify black people as having normal lung function despite having worse outcomes. When defining normality using LLN_white_ for all, people with normal FVC had similar breathlessness and mortality in both white and black people.

a) **Breathlessness**

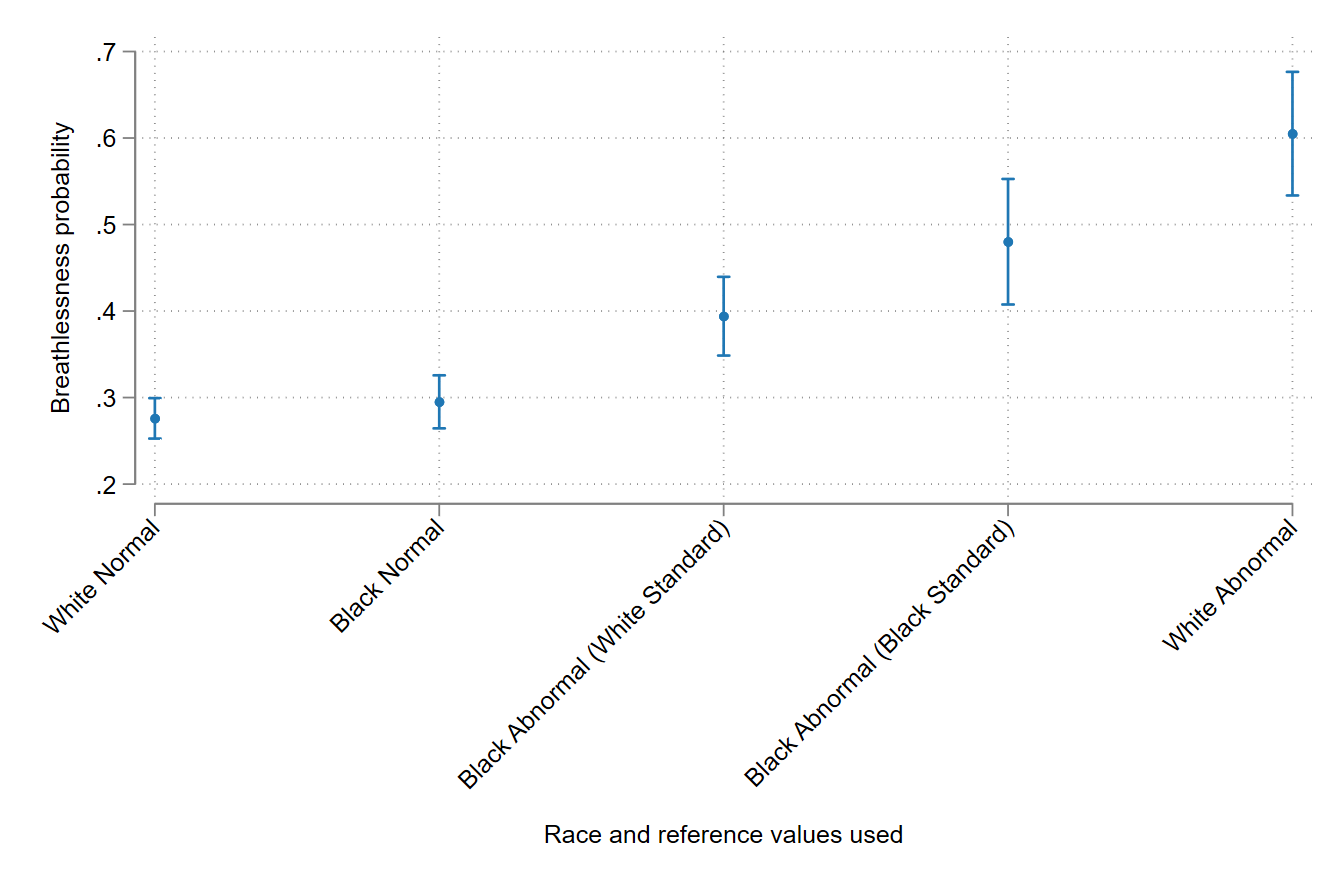

b) **Mortality**

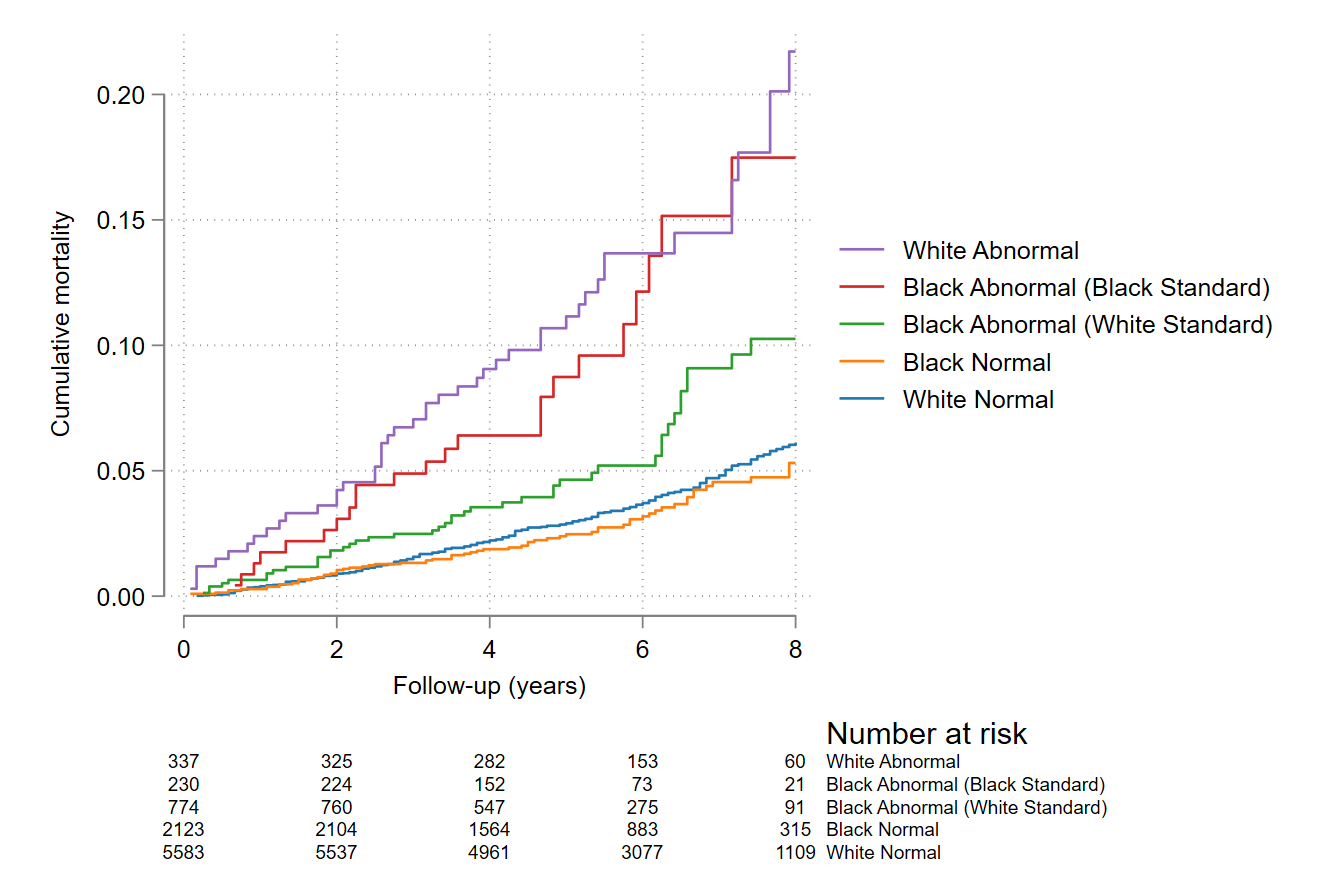
